# QRS prolongation is associated with structural remodeling in hypertrophic cardiomyopathy

**DOI:** 10.1101/2024.05.23.24307828

**Authors:** Hernan L. Vera-Sarmiento, Talha Tanriverdi, David Hurtado-de-Mendoza, Sanjay Sivalokanathan, Dolores Ketty, Daiyin Lu, Stefan Zimmerman, Sunil Sinha, Melvin Scheinman, M. Roselle Abraham

**Affiliations:** Hypertrophic Cardiomyopathy Center of Excellence, Division of Cardiology, University of California San Francisco, San Francisco, CA; Department of Medicine, SUNY Upstate Medical University, Syracuse, NY; Division of Cardiology, Johns Hopkins University School of Medicine, Baltimore Maryland; Cayetano Heredia University, Peru; Department of Radiology, Johns Hopkins University School of Medicine, Baltimore Maryland; Division of Cardiology, University of California San Francisco, San Francisco, CA

**Keywords:** hypertrophic cardiomyopathy, signal-averaged electrocardiogram, ventricular arrhythmias, myocardial fibrosis, left ventricular remodeling, global myopathy

## Abstract

**Background:** Signal-averaged electrocardiogram (SAECG) records myocardial depolarization, and can detect inhomogeneous/slow conduction in fibrotic myocardium, which promotes reentrant ventricular arrhythmias (VAs). Hypertrophic cardiomyopathy (HCM) is associated with a high prevalence of cardiac fibrosis and VAs, but abnormal SAECG has low predictive power for VAs. We hypothesized that HCM-specific structural/electrical remodeling underlies this result.

**Methods:** We tested our hypothesis by retrospectively studying HCM patients (n=73) who underwent transthoracic echocardiography (TTE) and cardiac magnetic resonance (CMR) imaging within 12 months of SAECG and 12-lead ECG. Patients were divided into 2 groups (normal-SAECG, abnormal-SAECG) based on filtered-QRS duration (*fQRS*d), root-mean-square-voltage (*RMS40*) and low-amplitude (<40μV) signal of terminal 40ms of filtered-QRS (late potentials). Abnormal SAECG was defined as *fQRS*d*>114ms, RMS40<20*μ*V* or *LAS40>38ms*.

**Results:** Abnormal SAECG was seen in ∼50% of HCM patients (37/73). In the abnormal-SAECG group, 78% (n=29) only had prolonged *fQRS*d, and 22% (n=8) had prolonged *fQRS*d plus late potentials (*RMS40<20*μ*V* or *LAS40>38ms)*. Mean *fQRS*d and *LAS40* were significantly higher in the abnormal-SAECG group. The abnormal-SAECG group had significantly larger LA size, lower global-LV longitudinal systolic strain/strain rate and early-diastolic strain rate by TTE; higher LV-mass index (LVMI) and LV-scar burden by CMR; higher prevalence of repolarization abnormalities on 12-lead ECG. LVEF and adverse outcomes (VT/VF, heart failure, death) were similar in the 2 groups. Univariate analysis showed that *fQRS*d is positively correlated with LVMI, LV-scar mass, and negatively correlated with global-LV early diastolic strain rate.

**Conclusions:** In HCM, abnormal SAECG is associated with greater structural/electrical LV-remodeling, reflecting a severe global myopathy.

## INTRODUCTION

Signal averaged electrocardiogram (SAECG) is a surface high-resolution technique that detects impulse propagation in both ventricles (reflected in the filtered QRS complex), as well as low-amplitude, high-frequency signals in the terminal 40ms of the QRS complex (labeled as late potentials) [1,2]. Late potentials result from slow and inhomogeneous impulse propagation in fibrotic myocardium which provides a substrate for reentrant ventricular arrhythmias [3,4]. Late potentials are common, and can predict risk for sustained ventricular arrhythmias (VAs), in patients with ischemic cardiomyopathy and arrhythmogenic right ventricular cardiomyopathy (ARVC) [5–8]. In contrast, hypertrophic cardiomyopathy (HCM), the most common cause of sudden death in young individuals, is also associated with a high prevalence of cardiac fibrosis, but SAECG has not been demonstrated to be useful for ventricular arrhythmia risk stratification in the general HCM population [9]. We hypothesized that HCM-specific structural/electrical remodeling and scar architecture underlies this result. Specifically, ischemic cardiomyopathy is associated with transmural scar in a vascular distribution, left ventricular (LV) dysfunction, eccentric LV remodeling that predisposes to anatomic reentry and inducible ventricular arrhythmias, whereas HCM hearts typically have patchy mid-myocardial scar, varying patterns of LV hypertrophy, hyperdynamic LV function and susceptibility to functional reentry without inducible ventricular arrhythmias [10].

In this retrospective study, we examined the relationship of SAECG with structural/electrical LV remodeling and adverse outcomes, in 73 HCM patients who underwent deep clinical phenotyping with SAECG, 12-lead ECG, rest/stress transthoracic echocardiography (TTE), cardiac magnetic resonance (CMR) imaging.

## METHODS

Electronic health records of all HCM patients who were evaluated at the Johns Hopkins HCM Center of Excellence (JH-HCM-COE) between 2009 and 2016 were retrospectively screened for SAECG and 12-lead ECG data. Hypertrophic cardiomyopathy is defined as left ventricular hypertrophy (LVH) with maximum wall thickness ≥15mm, and/or septal-to-posterior free wall ratio >1.3 by transthoracic echocardiography, in the absence of other causes such as hypertension and/or valvular disease [11]. Patients who underwent standard of care rest/stress TTE/ECG, contrast-enhanced cardiac magnetic resonance (CMR) imaging, and SAECG within a 12-month period were considered for inclusion in our study (**Figure 1**).

**Figure 1:**
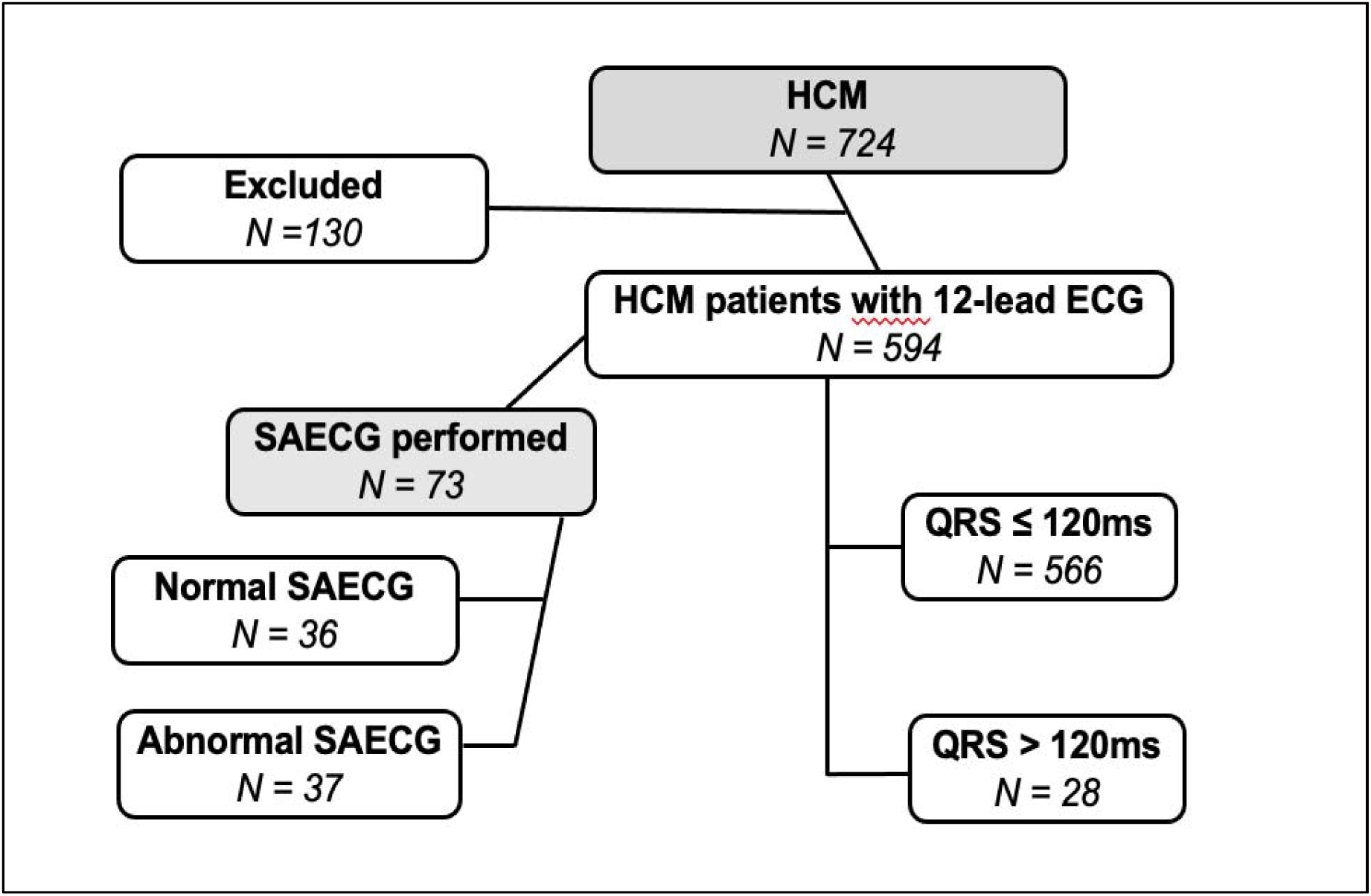
Flowchart of HCM patient selection. ECG: electrocardiogram, HCM: hypertrophic cardiomyopathy, SAECG: signal-averaged electrocardiogram.

The median follow-up was 2.1 (inter-quartile range 1.0-4.7) years for the overall cohort. Implantable cardioverter defibrillator (ICD) discharges, ventricular tachycardia/fibrillation (VT/VF) events were recorded by reviewing exercise ECG tracings, Holter, ICD interrogation reports, and clinic visit notes. Sustained VT was defined as VT at a rate ≥130 beats per minute and >30 sec duration, or VT that resulted in an ICD shock or anti-tachycardia pacing [12]. Heart failure (HF) was defined as new onset, or worsening HF to NYHA functional class III or IV requiring hospitalization. All-cause mortality statistics for our study population were obtained by linking our database to the Social Security Death Index.

### Study design (Figure 1)

We retrospectively identified 724 patients from the JH-HCM Registry who underwent SAECG, 12-lead ECG, TTE, and CMR within a year; of these, 130 patients were excluded due to history of myocardial infarction, alcohol septal ablation, septal myectomy, ECG evidence of bundle branch block or ventricular pacing, poor ECG traces, or incomplete CMR data. Of the remaining 594 patients, 73 had SAECG data, and all had 12-lead ECG data (**Figure 1**).

The following three values were recorded from SAECG data (**Figure 2**): (1) filtered QRS complex duration (*fQRS*d); (2) root-mean-square-voltage in the last 40ms of the filtered QRS (*RMS40*); (3) duration of the low-amplitude (<40μV) signal in the terminal filtered QRS (*LAS40*). Once recorded, each data point was assigned a threshold value that would differentiate it into a normal or abnormal result: *fQRS*d >114ms, *RMS40* <20μV, and *LAS40* >38ms were used to determine a value as abnormal. Patients were assigned to the abnormal-SAECG group if any of these 3 values were abnormal, or to the normal-SAECG group when all 3 values were normal.

**Figure 2:**
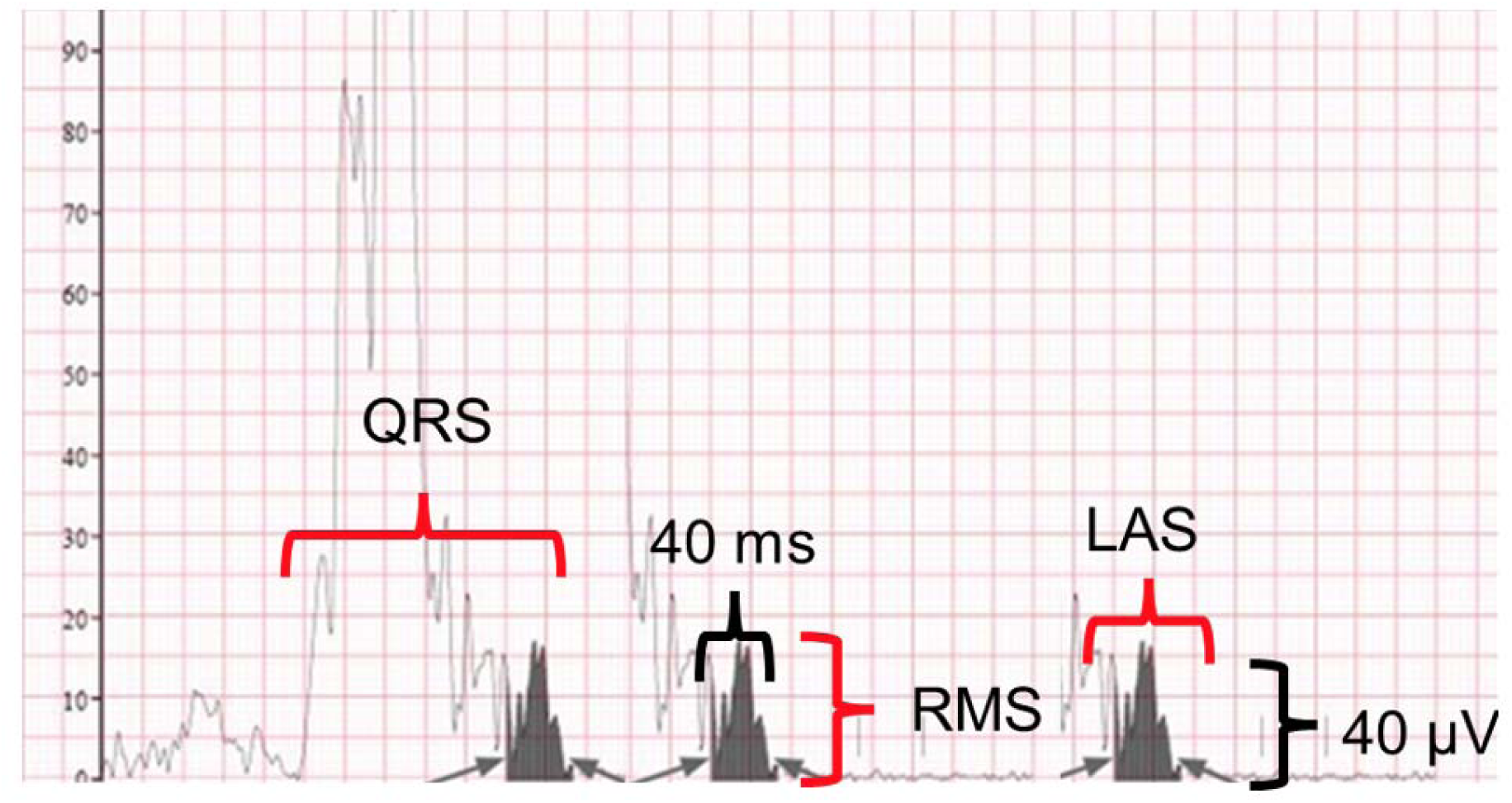
SAECG schematic.

### Data acquisition methods

#### Signal average electrocardiogram (SAECG) acquisition and analysis

Signal average ECG was recorded using a commercially available system (GE Marquette Mac 5500; GE Healthcare). Bipolar X, Y, and Z leads were used to record an average of 350 complexes. Each lead was filtered using a bidirectional filter at 40 to 250Hz. The three filter signals were then combined into a vector magnitude (X2+Y2+Z2)1/2. The noise level <0.4μV was set up for all cases.

#### Cardiac magnetic resonance image acquisition and analysis

*Image acquisition for assessment of cardiac morphology, function, replacement, and interstitial fibrosis:* Imaging was performed in a 1.5 Tesla system (MAGNETOM Avanto, Siemens Healthcare, Erlangen, Germany). The protocol included cine short-axis and four-chamber images with retrospective gating, using a torso coil array. Steady-state free precession (SSFP) images were obtained for cine four-chamber and short-axis images. For LGE quantification, images were obtained using 2D FLASH T1-weighted gradient-echo in short-axis view 10 min after administration of 0.2-mmol/kg gadopentetate dimeglumine. Myocardial T1 mapping was acquired with gradient echo multiphase-IR (TI scout) images from the Look-Locker (LL) sequence using a single image plane in the four-chamber view [13,14]. Sequence parameters included 17.92/1.09 TR/TE, 8-mm slice thickness, 50° flip angle, and 192 × 72 acquisition matrix.

*Evaluation of morphology, function, and replacement fibrosis in HCM hearts:* Epicardial and endocardial borders were manually contoured on the short-axis view in end-systole and end-diastole to calculate left ventricular mass, volume, function, and wall thickness using Qmass (version 7.4 Medis, Leiden, Netherlands). Replacement fibrosis was quantified by measuring late gadolinium enhancement (LGE) in the short-axis view. All LGE-positive images were analyzed quantitatively by manually contouring the LV endocardium and epicardium and excluding the papillary muscles. We used a semi-automated threshold of 6 standard deviations (SD) above the mean signal intensity of nulled myocardium to define LGE [15]. Areas of replacement fibrosis appear white, while the remainder of the myocardium is nulled and appears black (**Figure 3**). The extent of LGE is expressed as a percentage of total LV mass.

**Figure 3:**
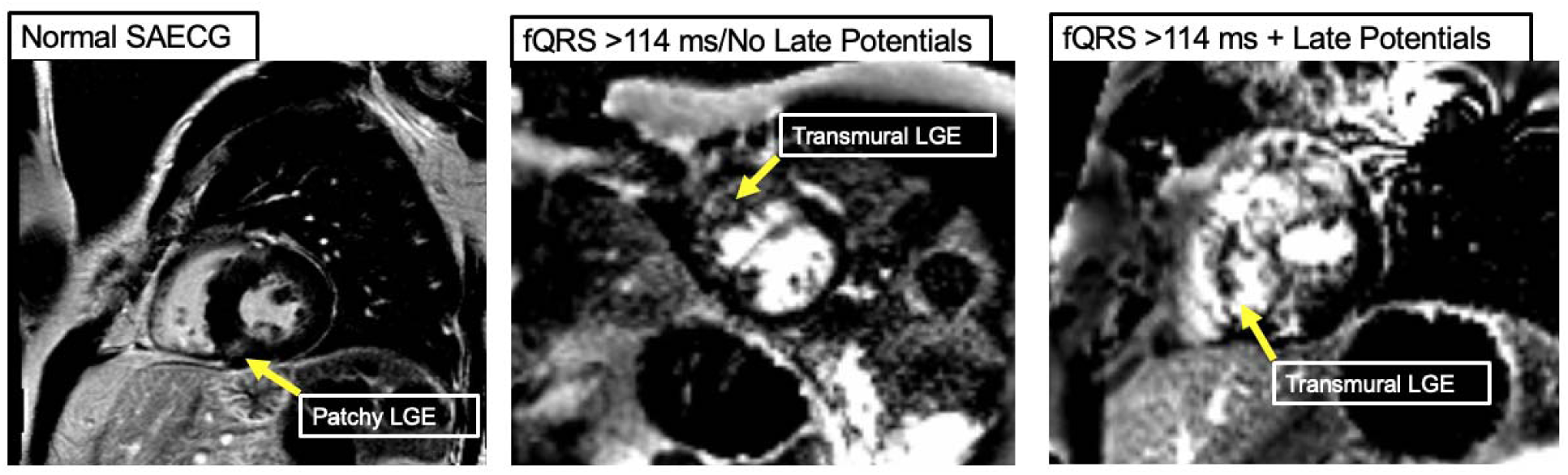
Late gadolinium enhancement patterns obtained by cardiac magnetic resonance imaging in HCM. Representative images of mid-myocardial LGE and transmural LGE in 3 HCM patients.

*Assessment of interstitial fibrosis in HCM hearts by T1 mapping*: For T1 mapping, the LL sequence was performed throughout the cardiac cycle, applying a 180° pulse and acquiring images at multiple inversion times to determine the null time of the normal myocardium. The LV endocardium and epicardium were manually contoured in all phases of the LL sequence in a four-chamber view, correcting for cardiac motion variations [10]. The mean myocardial T1 relaxation time of the LV was calculated using Mass Research software (version 5, Leiden, Netherlands). The mean T1 relaxation times were normalized to a standard gadolinium contrast dose, variation in delay times between contrast administration and sequence acquisition time, body surface area, and glomerular filtration rate, using a multi-compartment model [14].

#### Echocardiography image acquisition and analysis

*Assessment of cardiac morphology, function, hemodynamics by 2-D ECHO in HCM patients:* Transthoracic echocardiography was performed at baseline and following maximum treadmill exercise, as described previously [14]. HCM subtypes were defined, based on peak LV pressure gradients, measured at the level of the left ventricular outflow tract (LVOT) and mid-LV, at rest and following provocation (valsalva, exercise, amyl nitrite). HCM was classified as non-obstructive (gradients <30mmHg at rest and provocation), labile obstructive (rest gradients <30mmHg, provoked gradients ≥30mmHg), or obstructive (rest and provoked gradients ≥30mmHg) [16].

*Left ventricular deformation analysis in HCM patients*: Echocardiographic images for speckle tracking strain analysis were acquired at frame rates of 50-90Hz, and longitudinal strain was analyzed from the apical two-, three-, and four-chamber views using EchoPAC 112 (GE Vingmed Ultrasound AS, Horten, Norway). Peak longitudinal systolic and diastolic strain was measured in all 18 segments of the LV and subsequently averaged to obtain a global value, as described previously [17]. Poorly tracking segments or images that could not be optimized, were excluded from analysis. Patients with at least four unanalyzable segments were excluded from analysis [18].

#### Electrocardiographic (ECG) analyses

Standard 12-lead ECGs were obtained at rest, with patients in the supine position, and recorded at a paper speed of 25 mm/s. We selected ECGs that were recorded closest to the MRI date; heart rate, PR interval, QRS duration, QT interval, QTc (using Bazett’s formula) were measured automatically at acquisition and confirmed by two independent investigators (DH, TT) using Cardio Caliper Software (Iconico Inc), as described previously [12,13,17]. Pathological Q wave was defined as a Q wave with duration ≥0.03s, and/or amplitude ≥25% of the R wave amplitude, in at least two contiguous leads, in established lead groups. ST depression was defined as ≥0.5mm J point depression when compared to the PR segment. For males, ST elevation was defined as J point elevation ≥2mm in V2-V3, and ≥1mm in all other leads in patients >40 years old; in patients ≤40 years of age, J point elevation ≥2.5mm in V2-V3, and ≥1mm in all other leads was considered ST elevation. In females, ST elevation was defined as J point elevation ≥1.5mm in V2-V3, and ≥1mm in all other leads. Strain pattern was determined by the presence of contiguous ST-T inversion in the presence of prominent voltages in precordial leads [17]. Early repolarization pattern was defined by a notch or slur in the transition between the R wave and ST segment, J point peak ≥1mm in at least two anatomically contiguous leads, and QRS duration <120ms. QT intervals were measured from the QRS onset to the end of the T wave, defined as the intersecting point of a tangent line on the terminal T wave and TP baseline. Prolonged QTc interval was defined as ≥450ms in men, and ≥470ms in women, in the absence of bundle branch block or intraventricular conduction delay. Left bundle branch block (LBBB) was defined as QRS duration >120ms, QS pattern in V1, wide/tall R waves in V5-V6. Right bundle branch block (RBBB) was defined as QRS duration >120ms, rsr’, rsR’, or rSR’ pattern in leads V1 or V2 and broad S wave (≥40ms longer than R wave duration) in leads I, V6. QTc dispersion was defined as the difference between the maximum and minimum QTc in the following groups of leads that reflect electrical activity in septal/anterior (V1, V2, V3, V4), inferior (II, III, and aVF), and lateral (I, aVL, V5, and V6) walls.

#### Statistical analysis

Quantitative variables are presented using mean and standard deviation (SD). In the case of asymmetric distribution, variables are summarized by the median and the 25th and 75th percentiles. Student’s T-test was used to compare the mean between normal vs abnormal SAECG groups, after evaluating assumptions of normality and homogeneity of variances. If these assumptions were not met, non-parametric tests such as the U Mann-Whitney test were chosen. Qualitative variables are presented using frequencies. The Chi-square test was used to establish an association between these variables. Fisher’s exact test was used for expected frequencies of less than 5. Hypothesis tests were performed with a significance level (α) of 0.05, and the confidence intervals were estimated with a 95% confidence level. Correlation analyses were performed using Pearson’s and Spearman’s correlation coefficients. All procedures, from the preparation of the database for analysis, validation of the data, and statistical processing, were carried out using Stata, version 16 (StataCorp, College Station, TX).

## RESULTS

### Clinical characteristics of HCM patients stratified by SAECG data (**Table 1**)

Patients with SAECG data (n=73; **Figure 1**) were divided into 2 groups, namely, normal SAECG and abnormal SAECG based on duration of filtered QRS and presence of late potentials (**Table 1**). Mean patient age was similar in the two SAECG groups, but 78% of HCM patients in the abnormal SAECG group were male, compared to 47% in the normal SAECG group. HCM type, NYHA class, symptoms (excluding lightheadedness), comorbidities, and adverse outcomes were similar in the two groups. A similar proportion of patients in both groups were taking beta blockers and calcium channel blockers, but a greater number of patients in the abnormal SAECG group were prescribed angiotensin-converting enzyme (ACE) inhibitors or angiotensin receptor blockers; none of the patients were taking amiodarone.

**Table 1.** Clinical profile of HCM patients with Normal and Abnormal SAECG.

| Clinical features | Normal SAECC<br>(n=36) | Abnormal SAECC<br>(n=37) | p value |
| --- | --- | --- | --- |
| **Demographics** |  |  |  |
| Age (years) | 52.9 ± 13.7 | 49.9 ± 17.7 | 0.4 * |
| Male, n (%) | 17 (47.2) | 29 (78.4) | 0.008 ¥ |
| **HCM type** |  |  |  |
| Non-obstructive, n (%) | 11 (30.6) | 12 (32.4) | 1.0 ¥ |
| Labile obstructive, n (%) | 15 (41.7) | 16 (43.2) |  |
| Obstructive, n (%) | 9 (25.0) | 8 (21.6) |  |
| **Symptoms** |  |  |  |
| NYHA Class I, n (%) | 18 (50.0) | 21 (56.8) | 0.9 ¥ |
| NYHA Class II, n (%) | 11 (30.6) | 9 (24.3) |  |
| NYHA Class III, n (%) | 4 (11.1) | 5 (13.5) |  |
| Angina, n (%) | 14 (38.9) | 15 (40.5) | 1.0 ¥ |
| Dyspnea, n (%) | 19 (52.8) | 19 (51.4) | 1.0 ¥ |
| Syncope, n (%) | 7 (19.4) | 7 (18.9) | 1.0 ¥ |
| Lightheadedness, n (%) | 18 (50.0) | 8 (21.6) | 0.02 ¥ |
| **Comorbidity** |  |  |  |
| Diabetes, n (%) | 1 (2.8) | 2 (5.4) | 1.0 ¥ |
| Dyslipidemia, n (%) | 14 (38.9) | 17 (46.0) | 0.9 ¥ |
| Hypertension, n (%) | 18 (50.0) | 21 (56.8) | 0.9 ¥ |
| Coronary artery disease, n (%) | 3 (8.3) | 4 (10.8) | 1.0 ¥ |
| Myocardial infarction, n (%) | 0 (0.0) | 1 (2.7) | 1.0 ¥ |
| Stroke, n (%) | 0 (0.0) | 2 (5.4) | 0.6 ¥ |
| **Arrhythmia History** |  |  |  |
| NSVT, n (%) | 6 (16.7) | 5 (13.5) | 0.9 + |
| SVT, n (%) | 3 (8.3) | 6 (16.2) | 0.6 ¥ |
| Afib/flutter, n (%) | 3 (8.3) | 4 (10.8) | 1.0 ¥ |
| **Outcomes** |  |  |  |
| VT/VF, ICD shock, ATP (%) | 2 (5.5) | 0 (0) | 0.2 ¥ |
| New Afib/flutter (%) | 1 (2.7) | 1 (2.7) | 0.7 ¥ |
| Heart failure (%) | 1 (2.7) | 1 (2.7) | 0.7 ¥ |
| All-cause mortality (%) | 1 (2.7) | 0 (0) | 0.5 ¥ |
| **Medications** |  |  |  |
| Beta blocker, n (%) | 26 (72.2) | 23 (62.2) | 0.7 + |
| Ca-channel blocker, n (%) | 7 (19.4) | 7 (18.2) | 1.0 ¥ |
| ACE inhibitor/ARB, n (%) | 1 (2.8) | 13 (35.1) | 0.001 ¥ |
| Statin, n (%) | 11 (30.6) | 14 (37.8) | 0.6 ¥ |
| Diuretics, n (%) | 7 (19.4) | 10 (27.0) | 0.8 ¥ |
| * t-student test<br>+ Chi squared test<br>¥ Fisher Exact test<br>**Abbreviations:** **Afib:** atrial fibrillation, **ATP:** anti-tachycardia pacing, **ICD:** implantable cardioverter-defibrillator, **NSVT:** non-sustained ventricular tachycardia, **SVT:** supraventricular tachycardia, **VT/VF:** sustained ventricular tachycardia/ventricular fibrillation. |  |  |  |

### SAECG abnormalities in HCM patients

Abnormal SAECG, defined as *fQRS*d >114ms*, RMS40* <20μV or *LAS40* >38ms (**Figure 2**), was present in ∼50% (37/73) of HCM patients (**Table 2**). Majority of HCM patients with abnormal SAECG (n=29; 78%) demonstrated only prolonged filtered QRS duration (*fQRS*d >114ms); the remainder (n=8; 22%) had late potentials plus prolonged *fQRS*d. HCM patients in the abnormal SAECG group had significantly longer *fQRS* duration (*fQRS*d: 125.7±11.9 vs 107±4.7ms; p<0.0001), and low amplitude signal duration in the last 40 ms of the filtered QRS complex (*LAS40:* 31.2±13.9 vs 22.9±6 ms; p<0.0001) when compared to the normal SAECG group. No difference was observed in the root-mean-square voltage (*RMS40*) of the last 40 ms of the filtered QRS complex in the two groups.

**Table 2.**
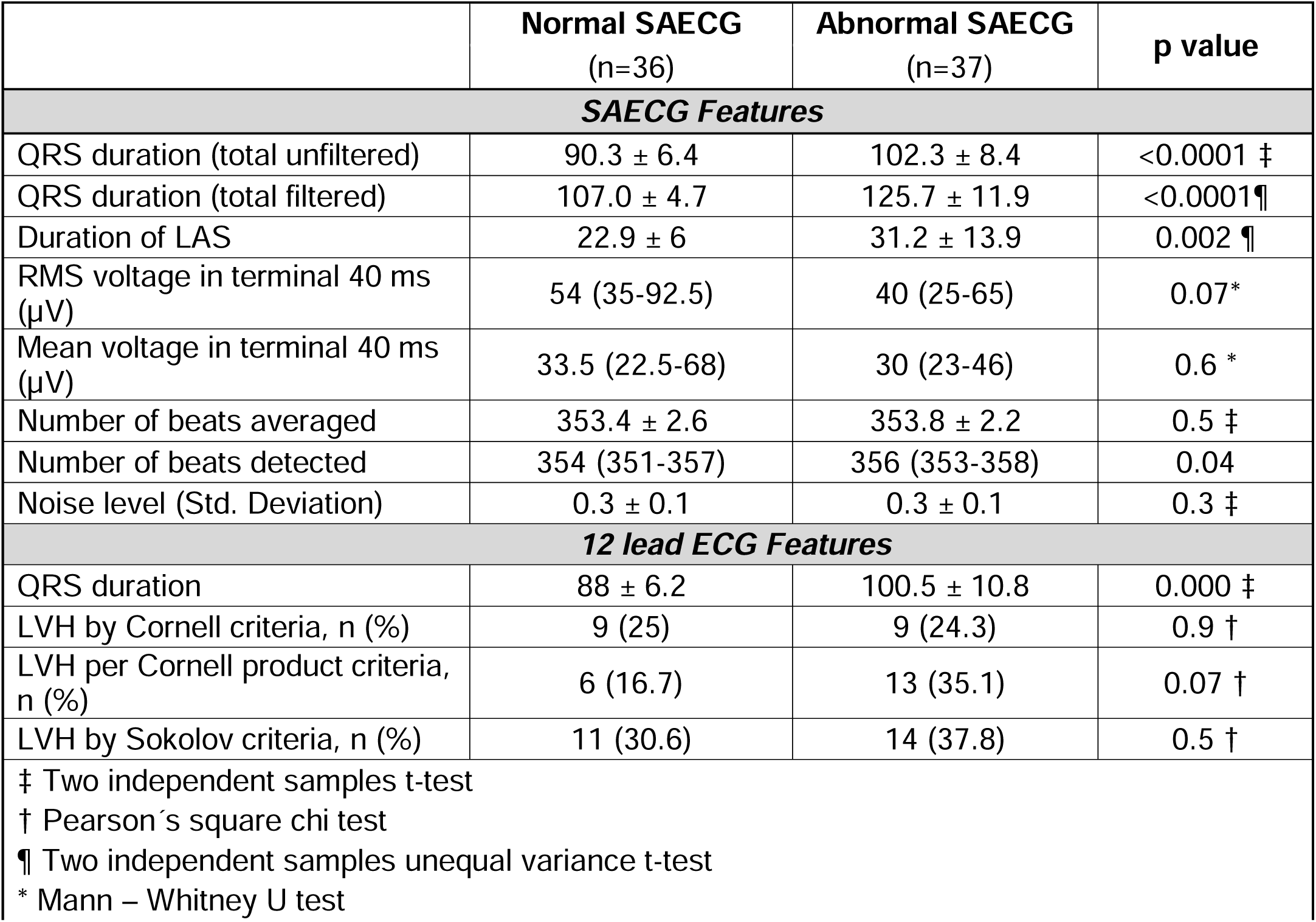
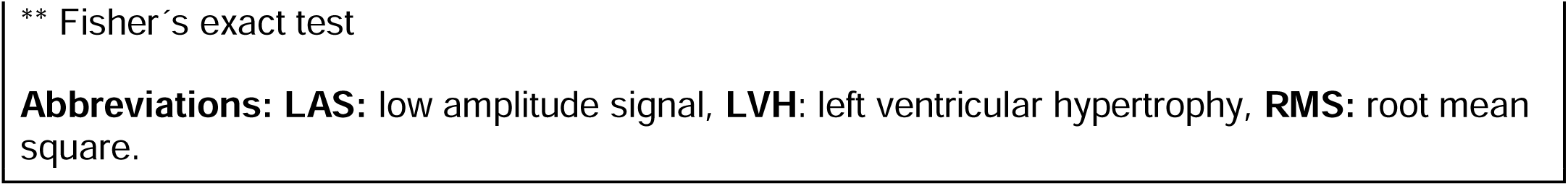
SAECG and 12-lead ECG features in HCM patients with Normal and Abnormal SAECG.

|  | **Normal SAECG**<br>(n=36) | **Abnormal SAECG**<br>(n=37) | **p value** |
| --- | --- | --- | --- |
| **SAECG Features** |  |  |  |
| QRS duration (total unfiltered) | 90.3 ± 6.4 | 102.3 ± 8.4 | <0.0001 ‡ |
| QRS duration (total filtered) | 107.0 ± 4.7 | 125.7 ± 11.9 | <0.0001 ¶ |
| Duration of LAS | 22.9 ± 6 | 31.2 ± 13.9 | 0.002 ¶ |
| RMS voltage in terminal 40 ms (µV) | 54 (35-92.5) | 40 (25-65) | 0.07* |
| Mean voltage in terminal 40 ms (µV) | 33.5 (22.5-68) | 30 (23-46) | 0.6 * |
| Number of beats averaged | 353.4 ± 2.6 | 353.8 ± 2.2 | 0.5 ‡ |
| Number of beats detected | 354 (351-357) | 356 (353-358) | 0.04 |
| Noise level (Std. Deviation) | 0.3 ± 0.1 | 0.3 ± 0.1 | 0.3 ‡ |
| **12 lead ECG Features** |  |  |  |
| QRS duration | 88 ± 6.2 | 100.5 ± 10.8 | 0.000 ‡ |
| LVH by Cornell criteria, n (%) | 9 (25) | 9 (24.3) | 0.9 † |
| LVH per Cornell product criteria, n (%) | 6 (16.7) | 13 (35.1) | 0.07 † |
| LVH by Sokolov criteria, n (%) | 11 (30.6) | 14 (37.8) | 0.5 † |
| ‡ Two independent samples t-test<br>† Pearson's square chi test<br>¶ Two independent samples unequal variance t-test<br>* Mann – Whitney U test |  |  |  |

\*\* Fisher's exact test
**Abbreviations:** **LAS:** low amplitude signal, **LVH:** left ventricular hypertrophy, **RMS:** root mean square.

### Cardiac HCM phenotype: imaging and ECG characteristics

*Echocardiography* (**Table 3**): Left ventricular ejection fraction, rest/provoked LV gradients, and diastolic function (E/A, E/e′) were similar in the two SAECG groups. However, the abnormal SAECG group had significantly higher left atrial (LA) diameter, lower peak global LV systolic strain/strain rate, and lower global LV early diastolic strain rate when compared to the normal SAECG group.

**Table 3.** Echocardiographic features of HCM patients with Normal and Abnormal SAECG.

|  | **Normal SAEKG**<br>(n=36) | **Abnormal SAEKG**<br>(n=37) | **p value** |
| --- | --- | --- | --- |
| LVEF (%) | 65.5 ± 8.2 | 64.9 ± 7.7 | 0.9 * |
| Max wall thickness (mm) | 2.0 ± 0.5 | 2.1 ± 0.4 | 0.4 * |
| Left atrial diameter (cm) | 3.9 ± 0.6 | 4.3 ± 0.8 | 0.03 * |
| LVEDV | 84 (68-115) | 91 (79-120) | 0.2 + |
| LVESV | 31.5 ± 11.1 | 36.6 ± 16.9 | 0.1 ** |
| Rest LVOTG (mmHg) | 16 (7-45) | 12.5 (9-26.5) | 0.7 + |
| Stress LVOTG (mmHg) | 48 (20-93) | 56 (24.5-103.5) | 0.6 + |
| E/A ratio | 1.3 ± 0.5 | 1.2 ± 0.4 | 0.5 * |
| E/e' ratio | 14 (12-18) | 16 (12-21) | 0.2 + |
| **Cardiac mechanics** |  |  |  |
| Global SG | -16.5 ± 3.1 | -14.8 ± 3.1 | 0.04 * |
| Global SRS | -1.0 ± 0.1 | -0.9 ± 0.2 | 0.008 * |
| Global SR E | 1.2 ± 0.3 | 1.0 ± 0.3 | 0.01 * |

\* t-student test
¥ Median (25<sup>th</sup> – 75<sup>th</sup> percentile)
+ U Mann Whitney Test
\*\* t-student test unequal
**Abbreviations:** **LVEF:** left ventricular ejection fraction, **LVEDV:** left ventricular end diastolic volume, **LVESV:** left ventricular end systolic volume, **LVOTG:** left ventricular outflow tract gradient, **SG:** peak systolic strain, **SR S:** peak systolic strain rate, **SR E:** early diastolic strain rate.

*Cardiac magnetic resonance imaging* (**Table 4**): HCM patients in the abnormal SAECG group had greater degree of LV hypertrophy, reflected by higher LV mass index (LVMI), than the normal SAECG group. Replacement fibrosis, which mainly consisted of mid-myocardial LGE, was seen in both groups, but the abnormal SAECG group had higher LV scar burden and higher prevalence of mid-myocardial scar when compared to the normal SAECG group. Myocardial T1 time, which reflects interstitial fibrosis, was similar in the two groups.

**Table 4.**
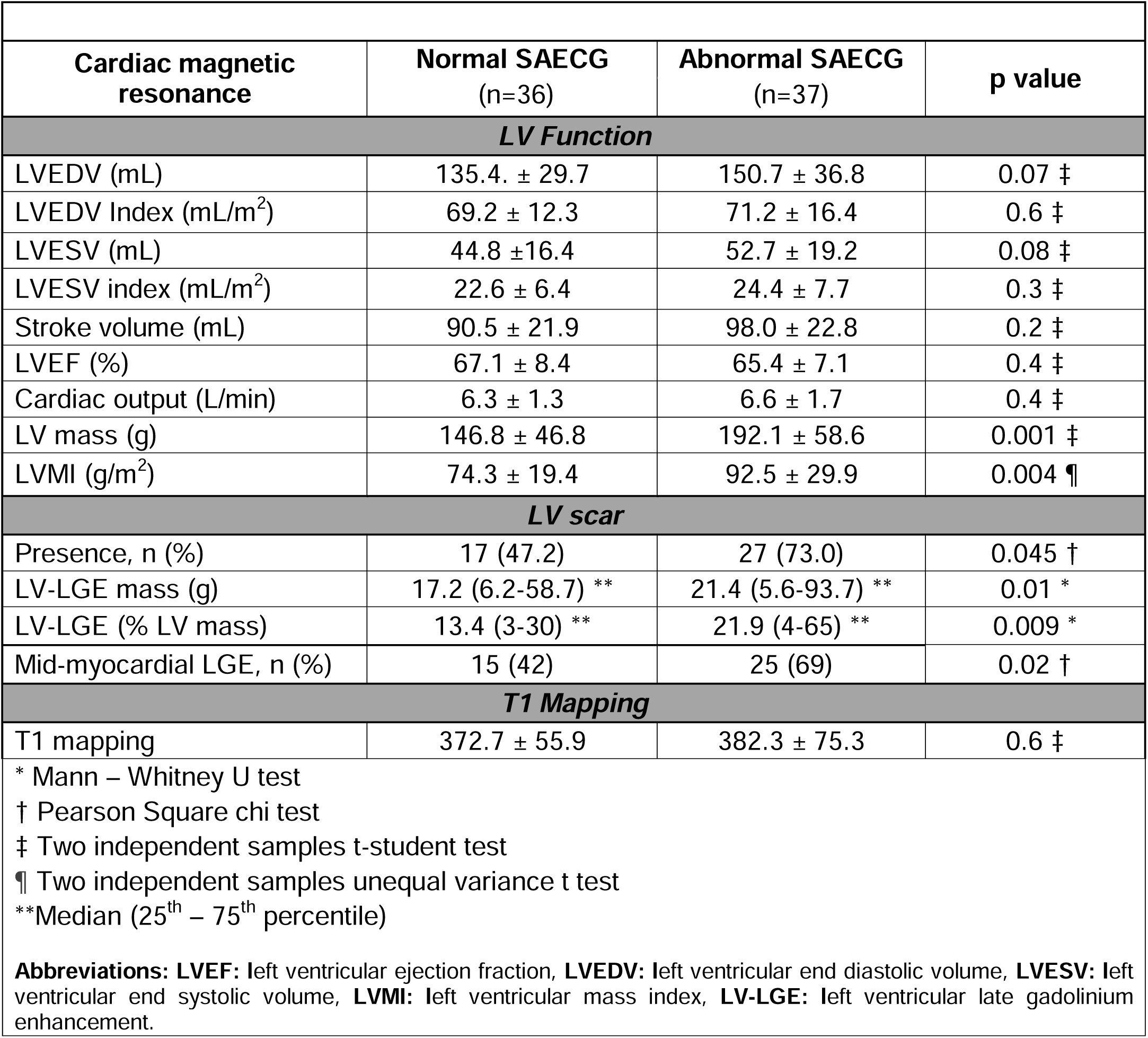
Cardiac magnetic resonance imaging features in HCM patients with Normal and Abnormal SAECG.

| Cardiac magnetic resonance | Normal SAECG<br>(n=36) | Abnormal SAECG<br>(n=37) | p value |
| --- | --- | --- | --- |
| **LV Function** |  |  |  |
| LVEDV (mL) | 135.4. ± 29.7 | 150.7 ± 36.8 | 0.07 ‡ |
| LVEDV Index (mL/m<sup>2</sup>) | 69.2 ± 12.3 | 71.2 ± 16.4 | 0.6 ‡ |
| LVESV (mL) | 44.8 ± 16.4 | 52.7 ± 19.2 | 0.08 ‡ |
| LVESV index (mL/m<sup>2</sup>) | 22.6 ± 6.4 | 24.4 ± 7.7 | 0.3 ‡ |
| Stroke volume (mL) | 90.5 ± 21.9 | 98.0 ± 22.8 | 0.2 ‡ |
| LVEF (%) | 67.1 ± 8.4 | 65.4 ± 7.1 | 0.4 ‡ |
| Cardiac output (L/min) | 6.3 ± 1.3 | 6.6 ± 1.7 | 0.4 ‡ |
| LV mass (g) | 146.8 ± 46.8 | 192.1 ± 58.6 | 0.001 ‡ |
| LVMI (g/m<sup>2</sup>) | 74.3 ± 19.4 | 92.5 ± 29.9 | 0.004 ¶ |
| **LV scar** |  |  |  |
| Presence, n (%) | 17 (47.2) | 27 (73.0) | 0.045 † |
| LV-LGE mass (g) | 17.2 (6.2-58.7) ** | 21.4 (5.6-93.7) ** | 0.01 * |
| LV-LGE (% LV mass) | 13.4 (3-30) ** | 21.9 (4-65) ** | 0.009 * |
| Mid-myocardial LGE, n (%) | 15 (42) | 25 (69) | 0.02 † |
| **T1 Mapping** |  |  |  |
| T1 mapping | 372.7 ± 55.9 | 382.3 ± 75.3 | 0.6 ‡ |
| * Mann – Whitney U test<br>† Pearson Square chi test<br>‡ Two independent samples t-student test<br>¶ Two independent samples unequal variance t test<br>**Median (25<sup>th</sup> – 75<sup>th</sup> percentile)<br>**Abbreviations:** LVEF: left ventricular ejection fraction, LVEDV: left ventricular end diastolic volume, LVESV: left ventricular end systolic volume, LVMI: left ventricular mass index, LV-LGE: left ventricular late gadolinium enhancement. |  |  |  |

In order to obtain mechanistic insights into the genesis of *fQRS* prolongation and late potentials in HCM hearts, we divided HCM patients who underwent SAECG into 3 groups (Group 1: normal SAECG; Group 2: *fQRS*d >114; Group 3: *fQRS*d >114 + late potentials), and analyzed the patterns of LV-LGE (**Figure 3**). In all 3 groups, the most common pattern was patchy mid-myocardial LGE located in maximally hypertrophied regions. A small number of patients in all 3 groups had evidence of subendocardial or transmural LGE: 8% in Group 1; 17% in Group 2; 25% in Group 3 (**Supplemental Table 1**). None of the patients had evidence of apical HCM or apical aneurysm [19].

*Electrocardiography* **(Supplemental Table 2, Figure 4**): A third of patients from both SAECG groups had prolonged QTc intervals at rest on 12-lead ECG. Patients in the abnormal SAECG group had a higher prevalence of repolarization abnormalities, namely T wave inversion in inferior leads, when compared to the normal SAECG group (p=0.02).

**Figure 4:**
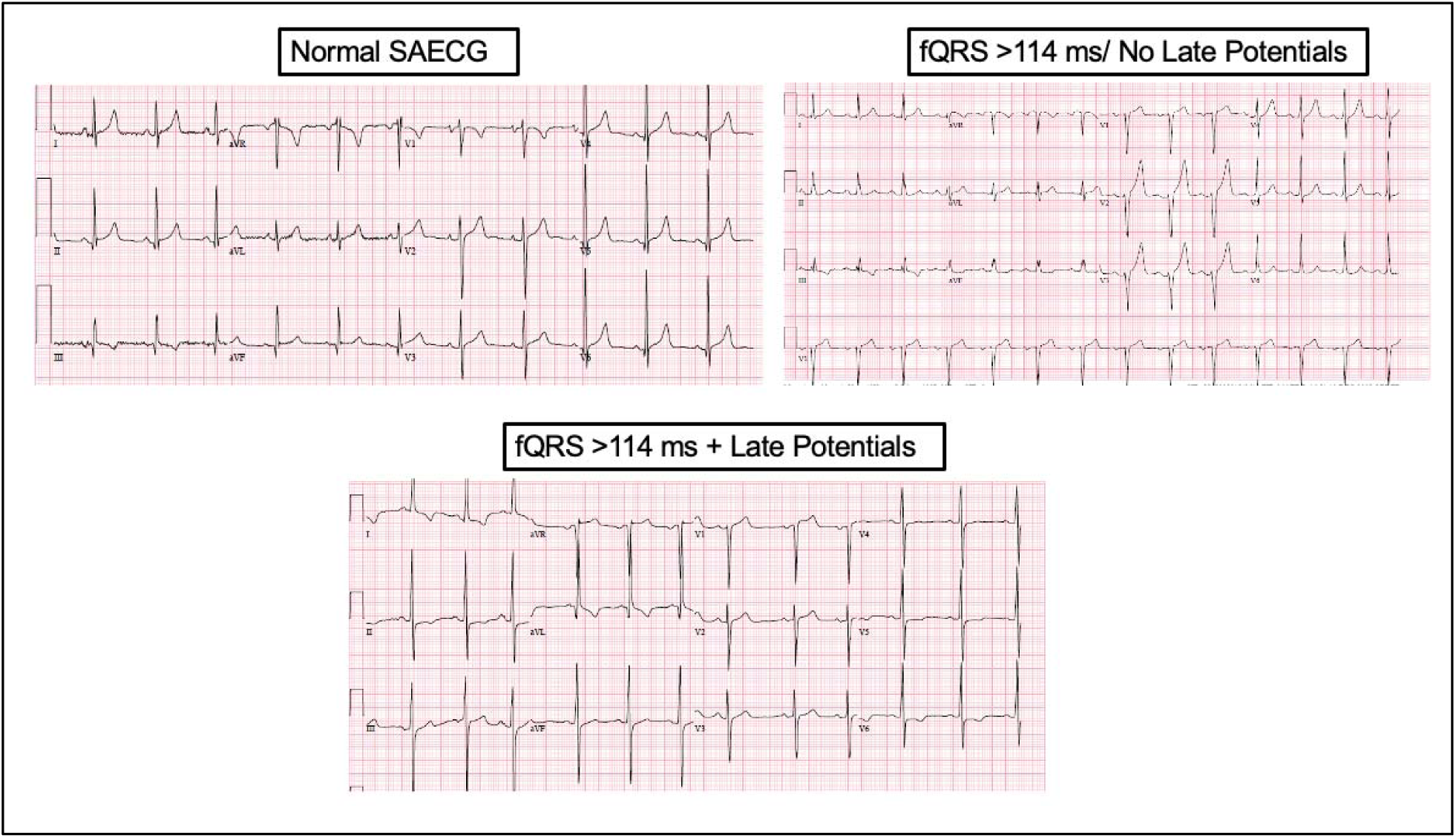
Representative ECGs in three HCM patients.

### Associations between SAECG and cardiac HCM phenotype (Table 5)

Since a large proportion (78%, n=29) of patients in the abnormal SAECG group had prolonged *fQRS* duration (**Figure 5A**), we examined the association between *fQRS* duration and cardiac morphology/function using univariate linear regression analyses (**Table 5**). We observed a statistically significant positive correlation between *fQRS* duration and LV mass index (r=0.43, p=0.0002; **Figure 5B**), LV scar mass (r=0.28, p=0.02), as well as a small negative correlation with global LV early diastolic strain rate (r=-0.34, p=0.009; **Figure 5C)**. As expected, a positive correlation was also observed between *fQRS* duration on SAECG and QRS duration on 12-lead ECG (r=0.67, p=0.00; **Table 5**, **Figure 6A**).

**Figure 5:**
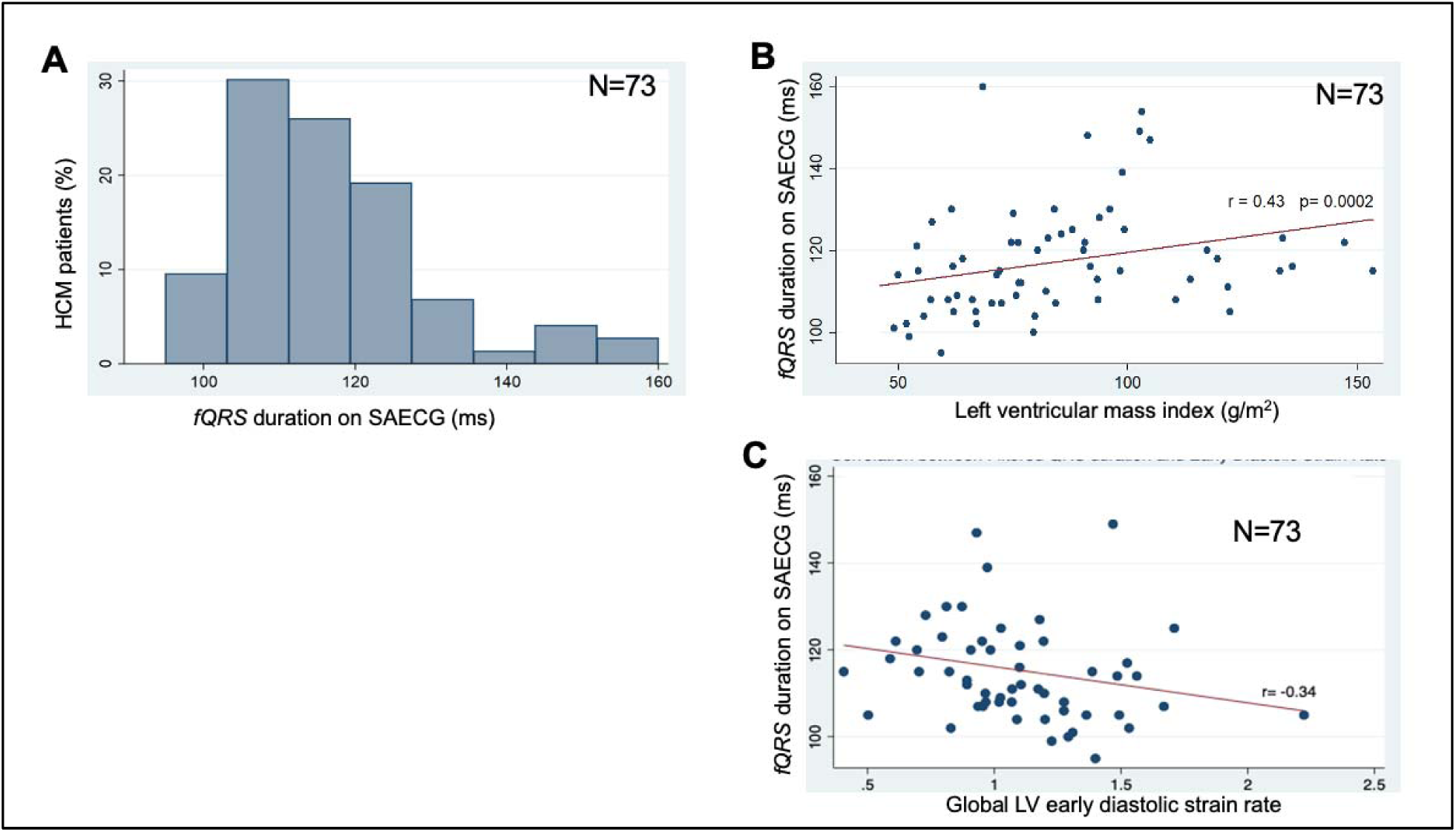
(A) Distribution of *fQRS* duration from SAECG, in HCM cohort (n=73). Positive correlation of *fQRS* duration with **(B)** left ventricular mass index. **(C)** inverse correlation with global LV early diastolic strain rate.

**Figure 6:**
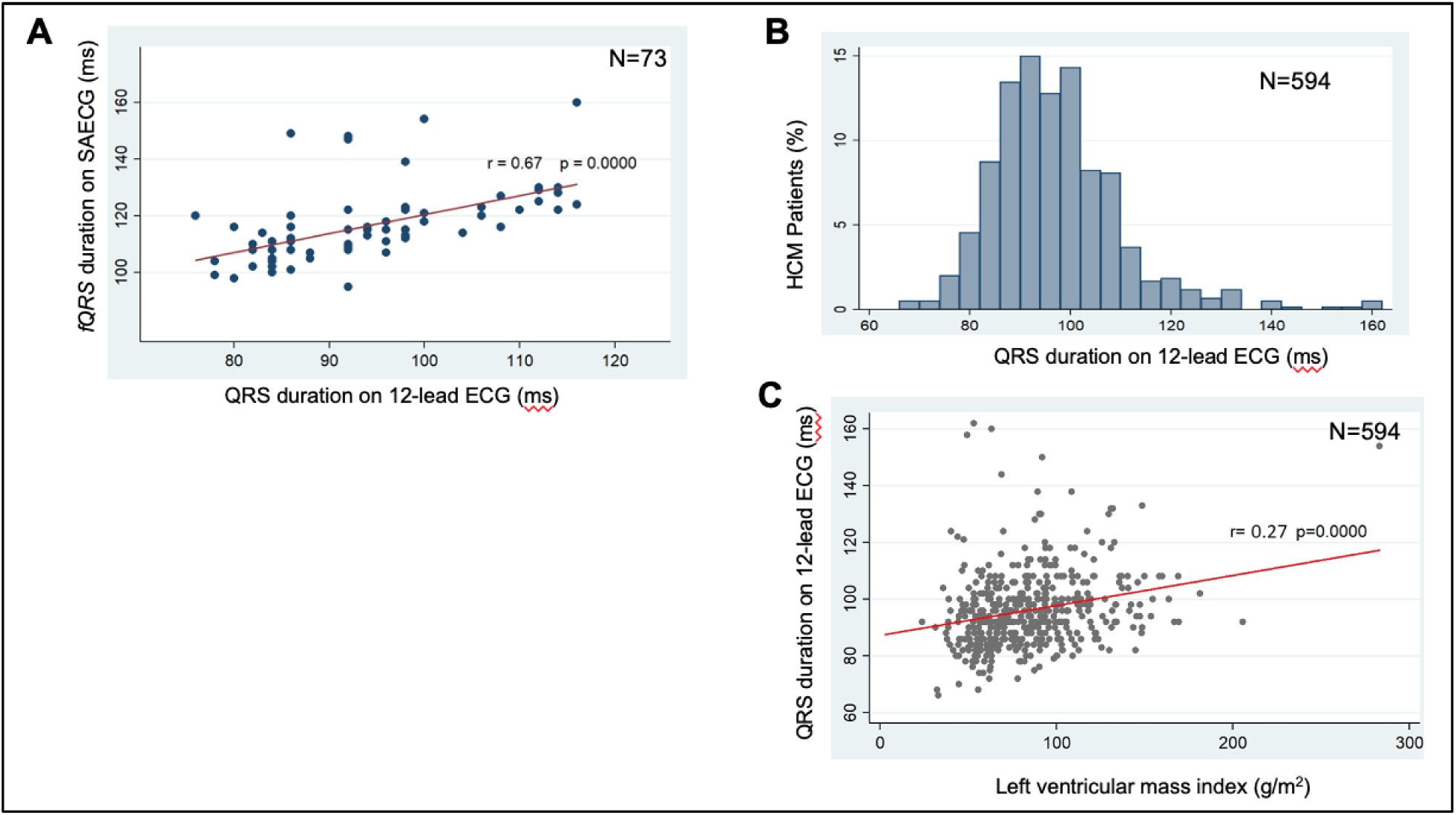
(A) Positive correlation between *fQRS* duration from SAECG and QRS duration from 12-lead ECG and **(B)** Distribution of QRS duration from 12-lead ECG in HCM cohort (n=594). **(C)** Positive correlation between QRS duration from 12-lead ECG and left ventricular mass index.

**Table 5.** Correlations between QRS duration (SAECG, 12-lead ECG) and imaging features in HCM.

| *fQRS* duration in SAECG | r value | p value | QRS duration on 12-lead ECG | r value | p value |
| --- | --- | --- | --- | --- | --- |
| LVMI | 0.43 | 0.0002 * | LVMI | 0.27 | 0.000* |
| LV mass | 0.39 | 0.001 * | LV Mass | 0.32 | 0.000* |
| LV scar tissue (g) | 0.28 | 0.02 * | LV scar tissue (g) | 0.09 | 0.03* |
| Global SG | 0.24 | 0.07 * | Global SG | 0.18 | 0.0002* |
| Global SR S | 0.24 | 0.07 * | Global SR S | 0.22 | 0.000* |
| Global SR E | - 0.34 | 0.009 * | Global SR E | -0.17 | 0.0008* |
| 12 Lead ECG QRSd (ms) | 0.67 | 0.0000 * |  |  |  |

\* Spearman correlation coefficient
**Abbreviations:** **LVMI:** left ventricular mass index; **SG:** peak systolic strain; **SR S:** peak systolic strain rate; **SR E:** early diastolic strain rate; **QRSd:** QRS complex duration

### QRS duration on 12 lead ECG is associated with structural remodeling

In order to confirm correlations between QRS duration and cardiac structure/function obtained from SAECG data (n=73), we went back to the entire JH-HCM cohort (n=724) who underwent deep clinical phenotyping (**Figure 1**). Patients with LBBB, RBBB, ventricular pacing, poor-quality ECGs (n=130) were excluded, resulting in the inclusion of 594 patients in this analysis (**Figure 1**).

The distribution of QRS duration in 12-lead ECG is presented in **Figure 6B** and shows that majority of HCM patients (n=566) had a 12-lead QRS duration ≤120 ms. Patients were then stratified into 2 groups using a QRS cutoff of 120 ms to examine differences in clinical characteristics: only LVMI and LA diameter were significantly different between the 2 groups (**Supplemental Table 3**) [9]. Next, we examined correlations between QRS duration on 12-lead ECG and cardiac structural/functional features (**Table 5**, **Figure 6C**): small but significant positive correlations were present between 12-lead QRS duration and LVMI (r=0.27, p=0.0), LV scar mass (r=0.09, p=0.03), global LV peak systolic strain (r=0.18. p=0.0002) and strain rate (p=0.22, p=0.00), as well as a small significant negative correlation with global LV early diastolic strain rate (r=-0.17, p=0.0008).

## DISCUSSION

In this study, we examined cardiac structural and electrical remodeling in HCM patients who underwent deep clinical phenotyping. Our results of a positive association between LV mass index and LV scar with QRS duration obtained by SAECG and 12-lead ECG suggest concomitant structural and electrical remodeling in HCM. The association of QRS duration with cardiac mechanics indicate that QRS prolongation could be a marker of worse myopathy in HCM patients.

Hypertrophic cardiomyopathy results from pathogenic variants in sarcomeric protein genes, and is the most common cause of sudden death in young individuals [20,21]. Expression of mutant sarcomeric proteins in cardiac myocytes leads to myocyte hypertrophy, interstitial and/or replacement fibrosis, microvascular and/or electrical remodeling. Increase in LV mass from myocyte hypertrophy is likely to be the main driver of high QRS voltage observed in 12-lead ECG, whereas changes in ion channel and/or gap junction expression/function in cardiac myocytes likely underlie QT prolongation and ST-T changes that are common in HCM [22].

Late potentials are microvolt oscillations in the terminal portion of the QRS complex and ST segment, that reflect heterogeneous and slow propagation in fibrotic myocardium [9]. Cardiac MRI in HCM patients has revealed a high prevalence of replacement fibrosis, which is typically patchy, and located in areas of maximal hypertrophy without associated regional wall motion abnormalities – this is in contrast to ischemic cardiomyopathy, where fibrosis is transmural, in a vascular territory and often associated with wall thinning as well as regional wall motion abnormalities. In ischemic cardiomyopathy, regions of infarct and the infarct border-zone are characterized by inhomogeneous and very slow impulse propagation, which manifest as late potentials on SAECG, and predispose to *anatomic reentry* (sustained monomorphic VT) [23]. In the case of HCM, global myocyte hypertrophy coupled with myocyte disarray, interstitial fibrosis, and/or gap junction remodeling would be expected to slow impulse propagation, manifesting as QRS prolongation in 12-lead ECG and SAECG. Slow, inhomogeneous impulse propagation in areas of patchy, replacement fibrosis could be hidden within the prolonged QRS, resulting in a low prevalence of late potentials on SAECG, despite a high prevalence of replacement fibrosis in HCM hearts. Electrical and structural remodeling in HCM could reduce wavelength of impulse propagation (by reducing conduction velocity) and cause wave-breaks, predisposing to *functional reentry* which can manifest as polymorphic VT or ventricular fibrillation [24]. The above differences in arrhythmic substrate between ischemic and hypertrophic cardiomyopathies could explain *1)* the low prevalence of late potentials, and poor predictive power of SAECG for ventricular arrhythmias in HCM, 2) easily induced sustained monomorphic VT in ischemic cardiomyopathy, but not in HCM patients with preserved LV function [19,25,26].

Prolongation of *fQRS >114ms* was the most common SAECG abnormality, and present in ∼50% of HCM patients in our study. We observed greater LV scar burden and higher prevalence of mid-myocardial LGE in patients with *fQRS* prolongation, and positive associations between LV mass index, LV scar mass and QRS duration on SAECG and 12-lead ECG (**Table 5**). This data suggests that cardiac fibrosis and hypertrophy contribute to *fQRS* prolongation in HCM. However, not all cardiac hypertrophy leads to prolonged *fQRS* duration: a prior SAECG study in healthy athletes with evidence of physiologic cardiac hypertrophy demonstrated a low prevalence (2.8%) of *fQRS* prolongation [27]. Taken together, our data suggests that QRS prolongation reflects pathologic cardiac hypertrophy and electrical remodeling in HCM.

## LIMITATIONS

This study has several limitations. The small number of patients who underwent SAECG and the short follow up time precludes a detailed assessment of the relationship between LV scar architecture, SAECG abnormalities and adverse outcomes such as ventricular arrhythmias and heart failure in HCM. This is a retrospective SAECG study that did not include patients with apical HCM and aneurysm, which could have increased the proportion of patients with late potentials on SAECG.

## CONCLUSION

Filtered QRS prolongation is the most common SAECG abnormality in HCM. QRS prolongation reflects pathologic hypertrophy and electrical remodeling, and could be useful as a marker of severe global myopathy in HCM.

## ETHICAL APPROVAL

The HCM Registries are approved by the Institutional Review Boards of the Johns Hopkins and the University of California San Francisco Hospitals. Informed consent was obtained for use of medical records for research.

## DISCLOSURE AND FUNDING

This study was supported by the JTB (John Taylor Babbit) foundation, and startup funds from the UCSF Division of Cardiology to MRA

## DECLARATION OF INTEREST

None

## Data Availability

All data produced in the present work are contained in the manuscript.

## Notes

### Competing Interest Statement

The authors have declared no competing interest.

