## Supplemental Tables for "QRS prolongation is associated with structural remodeling in hypertrophic cardiomyopathy"

**Supplemental Table 1. Cardiac HCM phenotype stratified by SAECG**

| **Cardiac HCM Phenotype (Cardiac Magnetic Resonance Imaging)** | **Normal SAECG**  **(*N=36*)** | **Abnormal SAECG (N=37)** | | **p value** |
| --- | --- | --- | --- | --- |
|  |  | ***fQRS* > 114**  **(*N=29*)** | ***fQRS*>114 + *RMS<20 + LAS40>38 (N=8)*** |  |
| LVMI (g) | 73 ± 21 | 95 ± 27 | 83 ± 19 | 0.003 * |
| LV mass (g) | 149 ± 48 | 194 ± 64 | 174 ± 47 | 0.007 * |
| Apical HCM or aneurysm, n (%) | 0 (0.0) | 0 (0.0) | 0 (0.0) |  |
| Mid-myocardial LGE | 15 (43) | 21 (72) | 4 (50) | 0.035 ^ |
| Subendocardial or transmural LGE, n (%) | 3 (8.3%) | 5 (17.2%) | 2 (25%) | 0.4 ^ |
| RV insertion LGE, n (%) | 2 (5.6%) | 0 | 1 (12.5%) | NS |
| Max wall thickness, cm | 1.99 ± 0.52 | 2.03 ± 0.57 | 1.96 ± 0.45 | 0.5 ** |
| LVEF, % | 67 ± 8 | 67 ± 7 | 65 ± 10 | 0.7 * |
| **Adverse outcomes** | | | | |
| VT/VF, ICD shock, ATP (%) | 2 (5.7) | 0 (0.0) | 0 (0.0) | 0.3 ^ |
| Atrial fibrillation/flutter | 1 (3.2) | 1 (4.16) | 0 (0.0) | 0.9 ^ |
| Heart failure (%) | 1 (3.2) | 0 (0.0) | 1 (14.2) | 0.2 ^ |
| All-cause mortality (%) | 1 (3.2) | 0 (0.0) | 0 (0.0) | 0.6 ^ |
| *ANOVA Test | | | | |
| **Kruskal-Wallis equality-of-populations rank test | | | | |
| ^ Chi squared Test | | | | |
| **Abbreviations: ATP**: anti-tachycardia pacing, **LGE:** late gadolinium enhancement, **LVMI: l**eft ventricular mass index, **LVEF:** left ventricular ejection fraction, **RV: r**ight ventricle, **VT/VF**: ventricular tachycardia / ventricular fibrillation. | | | | |

**Supplemental Table 2. 12-lead ECG features in HCM patients with Normal and Abnormal SAECG**

| **12-lead ECG Analysis** | **Normal SAECG** | **Abnormal SAECG** | **p value** |
| --- | --- | --- | --- |
|  | (n=36) | (n=37) |  |
| **QTc interval prolongation n (%)** | 11 (30.6) | 11 (29.7) | 0.9 † |
| **Pathologic Q waves** | 2 (5.6) | 8 (21.6) | 0.09 ** |
| I, aVL, V5-6 | 1 (2.8) | 5 (13.5) | 0.2 ** |
| V1-4 | 0 (0.0) | 1 (2.7) | 1.0 ** |
| II, III, aVF | 1 (2.8) | 2 (5.4) | 1.0 ** |
| **Right bundle branch block** | 0 | 0 |  |
| **Left bundle branch block** | 0 | 0 |  |
| **ST depression** | 8 (22.2) | 5 (13.5) | 0.3 † |
| I, aVL, V5-6 | 2 (5.6) | 1 (2.7) | 0.6 ** |
| V1-4 | 3 (8.3) | 3 (8.1) | 1.0 ** |
| II, III, aVF | 3 (8.3) | 1 (2.7) | 0.4 ** |
| **ST Elevation** | 1 (2.8) | 7 (18.9) | 0.06 ** |
| I, aVL, V5-6 | 0 (0.0) | 3 (8.1) | 0.2 ** |
| V1-4 | 1 (2.8) | 2 (5.4) | 1.0 ** |
| II, III, aVF | 0 (0.0) | 2 (5.4) | 0.5 ** |
| **T wave inversion** | 16 (44.4) | 35 (94.6) | 0.0 † |
| I, aVL, V5-6 | 8 (22.2) | 12(32.4) | 0.3 † |
| V1-4 | 5 (13.9) | 12 (32.4) | 0.06 † |
| II, III, aVF | 3 (8.3) | 11 (29.7) | 0.02 † |
| **Strain pattern** | 10 (27.8) | 18 (48.7) | 0.07 † |
| I, aVL, V5-6 | 6 (16.7) | 8 (21.6) | 0.6 † |
| V1-4 | 2 (5.6) | 5 (13.5) | 0.4 ** |
| II, III, aVF | 2 (5.6) | 5 (13.51) | 0.4 ** |
| **Early repolarization pattern** | 2 (5.6) | 1 (2.7) | 0.6 ** |
| I, aVL, V5-6 | 1 (2.8) | 0 (0.0) | 0.5 ** |
| V1-4 | 1 (2.8) | 1 (2.7) | 1.0 ** |
| II, III, aVF | 0 | 0 |  |
| ‡ Two independent samples t-test | | | |
| ¶ Two independent samples unequal variance t-test | | | |
| * Mann – Whitney U test | | | |
| ** Fisher´s exact test | | | |
| † Pearson´s square chi test | | | |

**Supplemental Table 3. Clinical characteristics of HCM patients stratified by QRS duration in 12-lead ECG**

| **Clinical features** | **QRS duration ≤ 120 ms** | **QRS duration > 120 ms** | **p value** |
| --- | --- | --- | --- |
|  | (n=566) | (n=28) |  |
| ***Demographics*** | | | |
| Age (years) | 53.4 ± 15.02 | 55.6 ± 20.1 | 0.46 * |
| Male, n (%) | 338 (59) | 19 (67) | 0.43 ¥ |
| ***HCM type*** | | | |
| Non-obstructive n, (%) | 190 (33.6) | 7 (25) | 0.49 ¥ |
| Labile obstructive, n (%) | 202 (35.7) | 9 (33) |  |
| Obstructive, n (%) | 173 (30.6) | 11 (40.7) |  |
| ***Symptoms*** | | | |
| NYHA Class I, n (%) | 275 (52) | 11 (42.3) | 0.52 ¥ |
| NYHA Class II, n (%) | 188 (35.6) | 11 (42.3) |  |
| NYHA Class III, n (%) | 61 (11.6) | 4 (15.3) |  |
| ***Arrhythmia History*** | | | |
| NSVT, n (%) | 50 (8.8) | 2 (7.1) | 0.7 + |
| VT/VF, n (%) | 14 (2.4) | 0 (0) | 0.5 ¥ |
| Atrial fibrillation/flutter, n (%) | 76 (13.4) | 5 (17.8) | 0.33 ¥ |
| ***Outcomes*** | | | |
| VT/VF, ICD Shock, ATP (%) | 17 (4.26) | 2 (9.52) | 0.24 ¥ |
| New AFib/flutter (%) | 30 (7.5) | 2 (9.5) | 0.48 ¥ |
| Heart failure (%) | 16 (4.0) | 0 (0.0) | 0.43 ¥ |
| All-cause mortality (%) | 9 (2.24) | 0 (0.0) | 0.62 ¥ |
| ***Medications*** | | | |
| Beta blocker, n (%) | 377 (66.6) | 20 (71.4) | 0.59 + |
| Calcium channel blocker, n (%) | 149 (26.3) | 11 (39.2)) | 0.10 ¥ |
| ***QRS duration*** | | | |
| QRS on 2-lead ECG (ms) | 94.5 ± 9.9 | 134 ± 12.3 | 0.0000 * |
| ***ECHO Imaging*** | | | |
| Max wall thickness (mm) | 2.0 ± 0.5 | 2.1 ± 0.6 | 0.11 * |
| Left atrial diameter (cm) | 4.1 ± 0.7 | 4.4 ± 0.6 | 0.04 * |
| Rest LVOTG (mmHg) | 28.2 ± 31.5 | 35.3 ± 31.2 | 0.28 * |
| Stress LVOTG (mmHg) | 68.3 ± 54.1 | 75.4 ± 63.6 | 0.52 * |
| E/A ratio | 1.32 ± 0.7 | 1.57 ± 1.61 | 0.13 * |
| E/e′ ratio | 16.4 ± 9.5 | 20.25 ± 12.8 | 0.04 * |
| ***LV Strain Imaging by ECHO*** | | | |
| Global SG | -16.12 ± 3.7 | -16.5 ± 3.6 | 0.62 * |
| Global SRS | -0.99 ± 0.2 | -1.04 ± 0.22 | 0.26 * |
| Global SR E | 1.13 ± 0.3 | 1.22 ± 0.4 | 0.28* |
| ***Cardiac Magnetic Resonance Imaging*** | | | |
| LVEF (%) | 69.6 ± 9.7 | 65.4 ± 14.4 | 0.05* |
| Mass (g) | 166.4 ± 64.7 | 185.2 ± 83.9 | 0.19 * |
| LVMI (g/m^2^) | 81.8 ± 29.5 | 96.8 ± 54.7 | 0.03 ¶ |
| LGE Presence, n (%) | 371 (70) | 22 (81.4) | 0.19 + |
| LGE mass (g) | 17.2 ± 26.19 | 19.5 ± 32.73 | 0.67 * |
| LGE (% of LV mass) | 9.6 ± 11.7 | 8.9 ± 10.4 | 0.79 * |
| * t-student test | | | |
| + Chi squared test | | | |
| ¥ Fisher Exact test | | | |
| ¶ Two independent samples unequal variance t test | | | |
| **Abbreviations**: **AFib**: atrial fibrillation; **ATP**: anti-tachycardia pacing; **NSVT**: non-sustained ventricular tachycardia; **VT/VF**: sustained ventricular tachycardia/ventricular fibrillation; **LVOTG:** left ventricular outflow tract gradient; **SG:** peak systolic strain; **SR S**: peak systolic strain rate; **SR E**: early diastolic strain rate; **LVEF:** left ventricular ejection fraction; **LVMI:** left ventricular mass index; **LGE:** late gadolinium enhancement | | | |
